# Identification of novel variants, genes and pathways potentially linked to Parkinson’s disease using machine learning

**DOI:** 10.1101/2023.06.20.23291658

**Authors:** Eric Yu, Roxanne Larivière, Rhalena A. Thomas, Lang Liu, Konstantin Senkevich, Shady Rahayel, Jean-François Trempe, Edward A. Fon, Ziv Gan-Or

## Abstract

There are 78 loci associated with Parkinson’s disease (PD) in the most recent genome-wide association study (GWAS), yet the specific genes driving these associations are mostly unknown. Herein, we aimed to nominate the top candidate gene from each PD locus, and identify variants and pathways potentially involved in PD. We trained a machine learning model to predict PD-associated genes from GWAS loci using genomic, transcriptomic, and epigenomic data from brain tissues and dopaminergic neurons. We nominated candidate genes in each locus, identified novel pathways potentially involved in PD, such as the inositol phosphate biosynthetic pathway (*INPP5F*, *IP6K2*, *ITPKB, PPIP5K2*). Specific common coding variants in *SPNS1* and *MLX* may be involved in PD, and burden tests of rare variants further support that *CNIP3*, *LSM7*, *NUCKS1* and the polyol/inositol phosphate biosynthetic pathway are associated with PD. Functional studies are needed to further analyze the involvements of these genes and pathways in PD.

## Introduction

Genome-wide association studies (GWAS) have nominated many variants associated with complex traits. In Parkinson’s disease (PD), the most recent GWAS revealed 90 independent risk variants across 78 genomic loci. ^1^ Although many single-nucleotide polymorphisms (SNPs) are in novel genomic loci, well-established PD genes discovered many years ago, such as *LRRK2*, *PINK1*, *DJ-1*, *SNCA*, *GBA1*, *PRKN* and *MAPT* (Supplementary Table 1) still account for the vast majority of research on Parkinson’s disease.

Several disadvantages of GWAS limit additional functional analyses. First, above 90% of all GWAS significant SNPs are in noncoding regions.^2^ These SNPs are often passenger variants due to complex linkage disequilibrium (LD). Second, the causal gene associated with the causal SNPs remains unclear in most GWAS loci.^3^ To overcome these challenges, downstream GWAS analyses were established with the aim of identifying causal genes within GWAS loci. This involves techniques such as fine-mapping and colocalization methods to nominate causal SNPs, as well as transcriptome-wide association studies to nominate gene-trait associations.^4–6^ These models use LD structure, and gene expression panels to discover causal SNPs/genes. While these methods may propose causal variants and genes, additional biological evidence is generally required to pair causal variants with causal genes. Using multi-omic analyses, one can integrate a diverse range of comprehensive cellular and biological datasets such as genomic, transcriptomic and epigenetic datasets and use platforms such as *Open Targets Genetics* (https://genetics.opentargets.org/) to perform systematic analyses of gene prioritization across all publicly available GWASs.^7^ Although powerful, *Open Targets Genetics* lacks disease-specific tissues relevant to PD such as dopaminergic neurons and microglia. Using a similar approach, we may discover additional pathways and genetic targets involved in PD.

In this study, we leveraged PD-relevant transcriptomic, epigenomic and other datasets in our gradient boosting model (Figure 1). We trained this model on well-established PD genes to nominate causal genes from PD GWAS loci.

**Figure 1:**
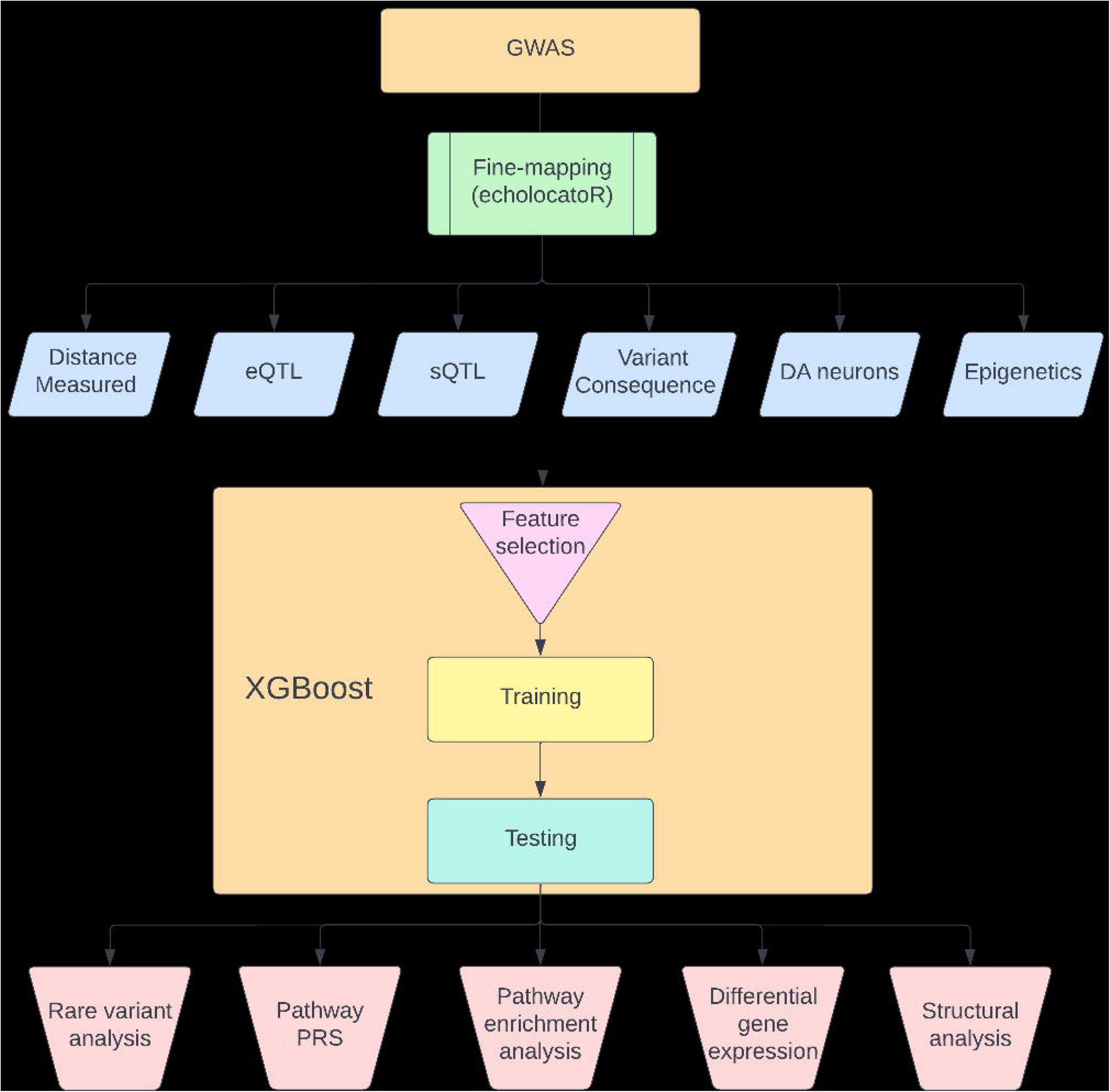
Workflow summary. This figure describes the analyses performed in this study.

## Results

### Machine learning model nominates PD-associated genes in each PD locus

To train our machine learning model, we used seven well-established PD-associated genes from the PD GWAS (*GBA1*, *LRRK2, SNCA, GCH1*, *MAPT*, *TMEM175*, *VPS13C*) as positive labels, and the remaining genes from the same loci (n=205) were used as negative labels (i.e. genes that are unlikely to be involved in PD). We trained an XGBoost regression model to identify the best predictive features. Then, based the best predictive features, we assigned a probability score that indicated the likelihood that the gene was driving the association at each locus (Supplementary Table 2). We then nominated the top-scoring genes in each locus (Supplementary Table 2, Figure 2). Two genes, *MAPT* and *TOX3*, were nominated twice in neighboring loci that harbor them, bringing the total number of genes nominated in this model to 76 genes in 78 loci. 48 of the 76 genes (63%) had a probability score higher than 0.75. Of note, five genes (*NEK1*, *FDFT1*, *PSD*, *BAG3* and *SLC2A13*) that were ranked second in their respective loci also had a probability score >0.75. However, the nominated genes in their loci (*CLCN3*, *CTSB*, *GBF1*, *INPP5F* and *LRRK2*, respectively) all had probability scores >0.94. In seven other loci, the top nominated genes had an especially low probability score (<0.3), including *RBMS3*, *HIST1H2BL*, *TRIM40*, *EHMT2*, *RPS12*, *MICU3* and *ITGA8*.

**Figure 2:**
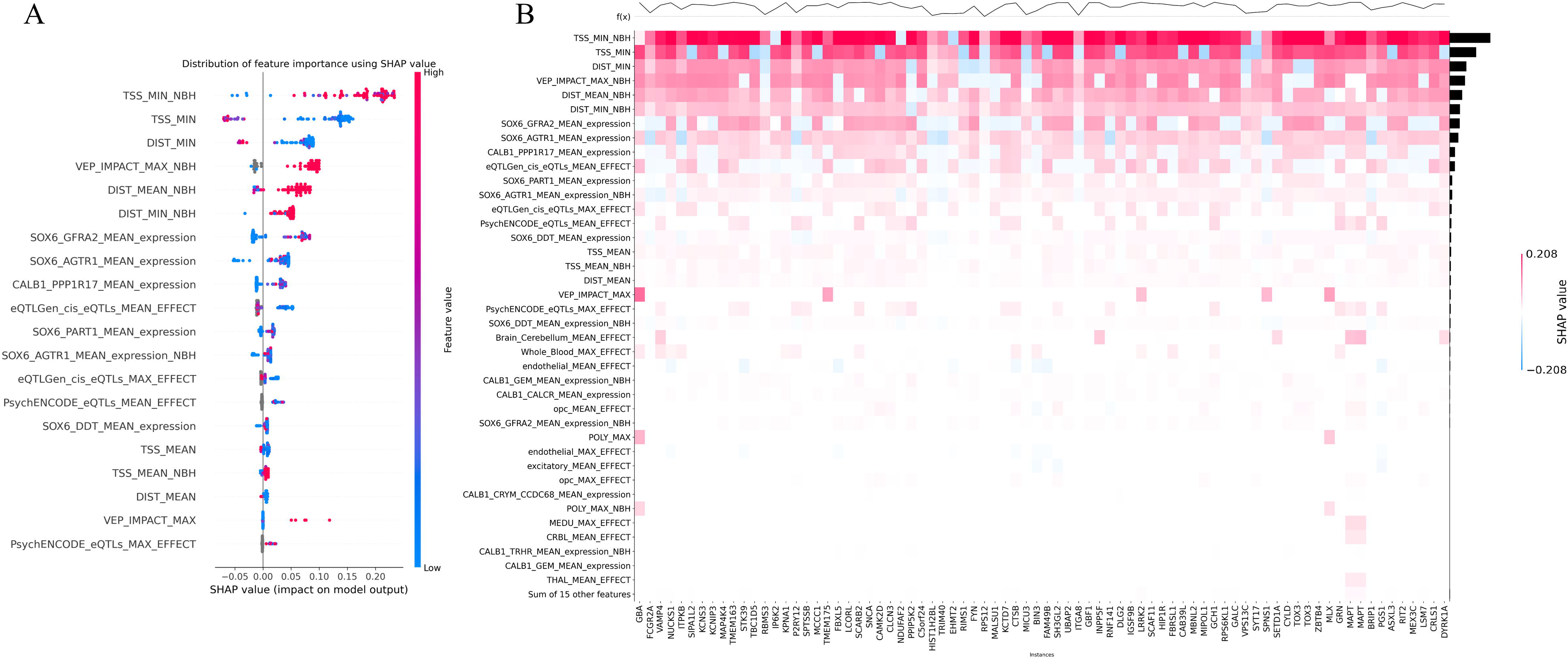
Probability score of the Parkinson’s disease GWAS candidate genes. Manahattan plot showing the probability scores from the machine learning model for each locus in the Parkinson’s disease GWAS loci sorted in descending order. For each gene, the top non-distance feature was used to color the data.

### Gene expression in specific subtypes of PD-associated dopaminergic neuron is an important feature predicting PD-relevant genes

Next, we used Shapley Additive exPlanations (SHAP) values to determine which features of the model contributed most to the prediction.^8,9^ SHAP values provide, for each gene, the relative contribution of each feature to the selection of that gene. The most important features for the scoring of each gene are shown in Figure 3A. As expected, distance-related features, such as distance from the top-associated SNP in the locus to the transcription start site or distance to the beginning of the gene, were the most important features in our model.^7^ The next most important feature was the Variant Effect Predictor (VEP) value, followed by additional distance measures.^10^ Interestingly, the next most important features were mRNA expression values within specific dopaminergic neuron subtypes. These different dopaminergic neuron subtypes are defined by the expression of the genes *GFRA2* and *AGTR1* from single nuclear sequencing of postmortem tissue. The latter is a specific subtype of dopaminergic neurons shown by Kamath et al. to be selectively degenerated in brains of PD patients.^11^ The remaining features include expression in other dopaminergic neuron subpopulation by Kamath et al, expression quantitative trait loci (eQTLs) and others expression features. Epigenetic features were not predictive in our model. As shown in Figure 3B, all nominated genes had at least one of the distance features contributing to their selection. On top of the known contribution of missense variants in *GBA1*, *LRRK2* and *GCH1*, we nominated missense SNPs that contributed to the score of two candidate genes: *SPNS1* (p.L512M, rs7140) and *MLX* (p.Q139R, rs665268). *SPNS1* and *MLX* have not been previously implicated in PD, and the important features identifying these genes as the top candidate for their respective GWAS loci are shown in Figure 4.

**Figure 3:**
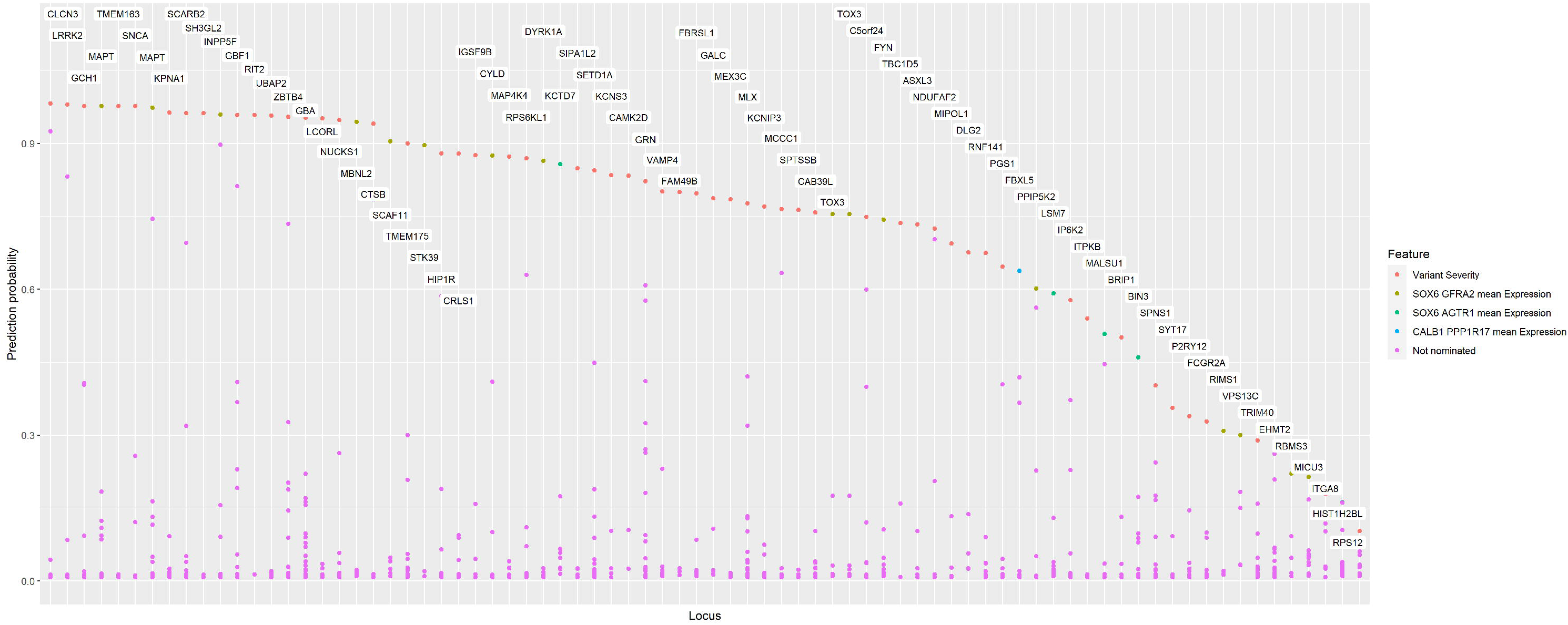
Feature importance for the Parkinson’s disease GWAS gene prioritization model. A) Bee-swarm plot of feature importance using SHAP values along with the distribution of genes based on feature value B) Heatmap of feature importance using SHAP value for the top candidate gene in each locus. The plot at the top represents the probability score of each gene. The bar plot on the right shows the relative importance of each feature.

**Figure 4:**
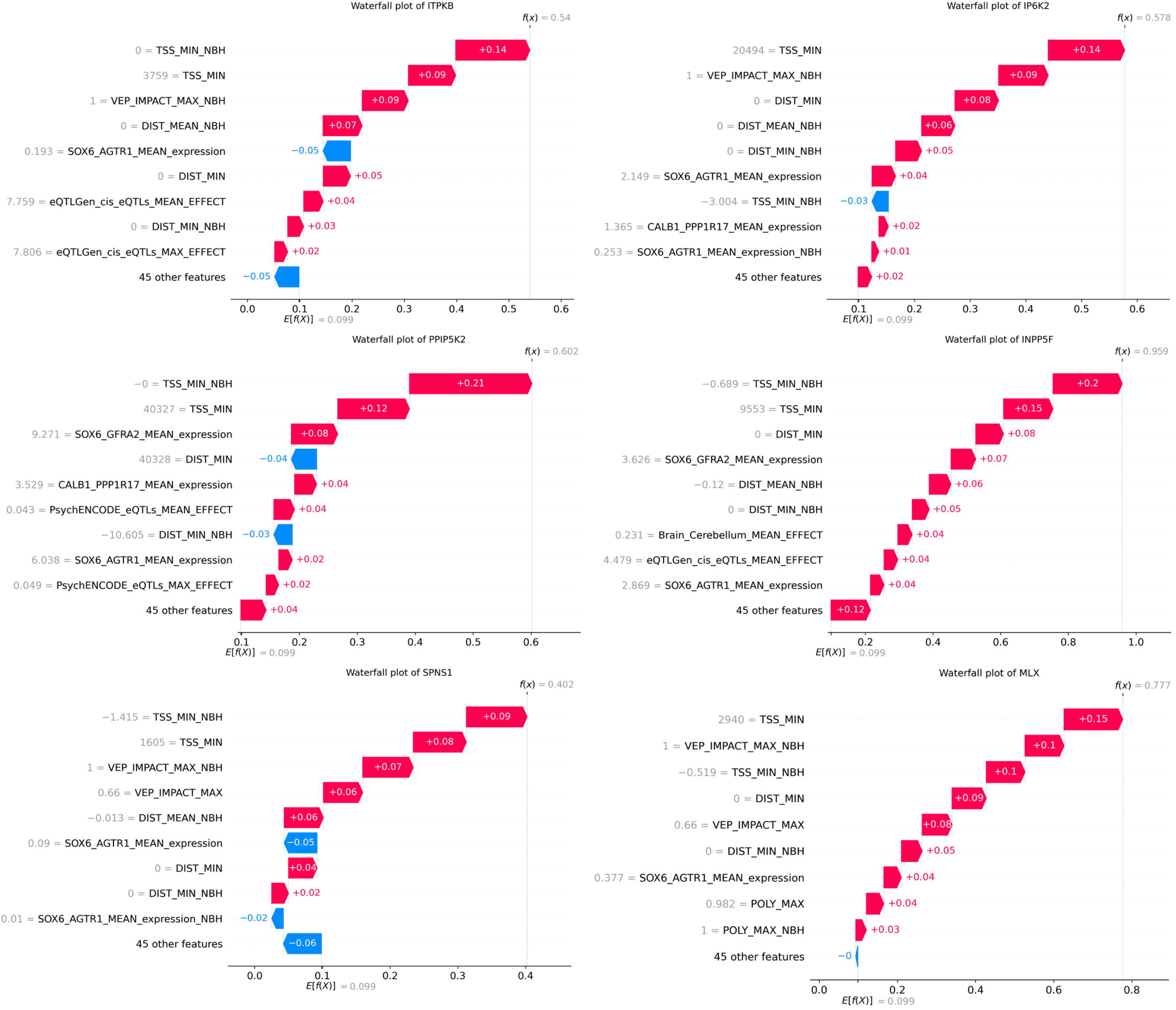
Waterfall plots for Parkinson’s disease GWAS candidate genes. Importance of the top 10 features using SHAP values for different selected candidate genes. E[f(x)] is the base score for each gene which is calculated based on the average value of each features. f(x) is the final score after accounting for all features.

### Differential gene expression of genes from the inositol phosphate biosynthetic pathway and *MLX1* in PD

To further establish the importance of the nominated genes in PD, we examined whether they are differentially expressed in PD patients compared to controls, using expression data from single nuclear RNAseq (scRNA) from Kamath et al^11^ and single nuclear and bulk RNAseq datasets from FOUNDIN-PD.^12^ Out of the top nominated genes, *INPP5F* (average log fold change[FC]LJ=LJ−7.22, *p*LJ=LJ2.90e-31) and *MLX* (average log FCLJ=LJ-1.80, *p*LJ=LJ2.23e-4) were associated with PD in the data from Kamath et al (Supplementary Table 3).^11^ In FOUNDIN-PD^12^, after excluding prodromal cases, we found differential expression of across many genes including *INPP5F* (average log FC =LJ0.070, *p*LJ=LJ1.89e-19) and *IP6K2* (average log FCLJ=LJ−0.076, *p*LJ=LJ1.35e-35) in scRNA data (n=80) from dopaminergic neurons by comparing PD and control (Supplementary Table 4). Results from the bulk RNAseq analysis (n=92) can be found in Supplementary Table 5.

### Structural analysis of *SPNS1* and *MLX*

Since nonsynonymous variants in *SPNS1* and *MLX* were identified as major contributors to their selection as the nominated genes in their loci, we aimed to examine the potential consequences of these variants by performing *in silico* structural analyses of the protein encoded by these genes. *SPNS1* encodes a transporter for phospholipids at the lysosome membrane.^13^ It mediates the efflux of lysophosphatidylcholine and lysophosphatidylethanolamine out of the lysosome. The SNP rs7140 is located in the 3’-untranslated region (UTR) of the canonical splice variant 1 transcript, which produces the 528 a.a. isoform that has been investigated functionally^13^ (Uniprot #Q9H2V7). This canonical isoform has also been observed in numerous proteomics datasets in gpmDB (https://gpmdb.thegpm.org/index.html). However, six other potential isoforms generated by alternative splicing have been predicted, including a 538 a.a. fragment with an alternative C-terminus where as the rs7140 SNP is located within the coding region (Uniprot #H3BR82). The rs7140 variant results in the p.L512M mutation in this isoform. To investigate the impact of this mutation on the function of this *SNPS1* isoform, we inspected the 3D structure model generated by AlphaFold.^14^ Leu512 is located in the unstructured C-terminus of this membrane-bound protein, on the lumenal side of the lysosomal membrane (Figure 5A). The role of the C-terminus in this isoform of *SPNS1* remains unclear, and thus the impact of the p.L512M mutation is unknown.

**Figure 5:**
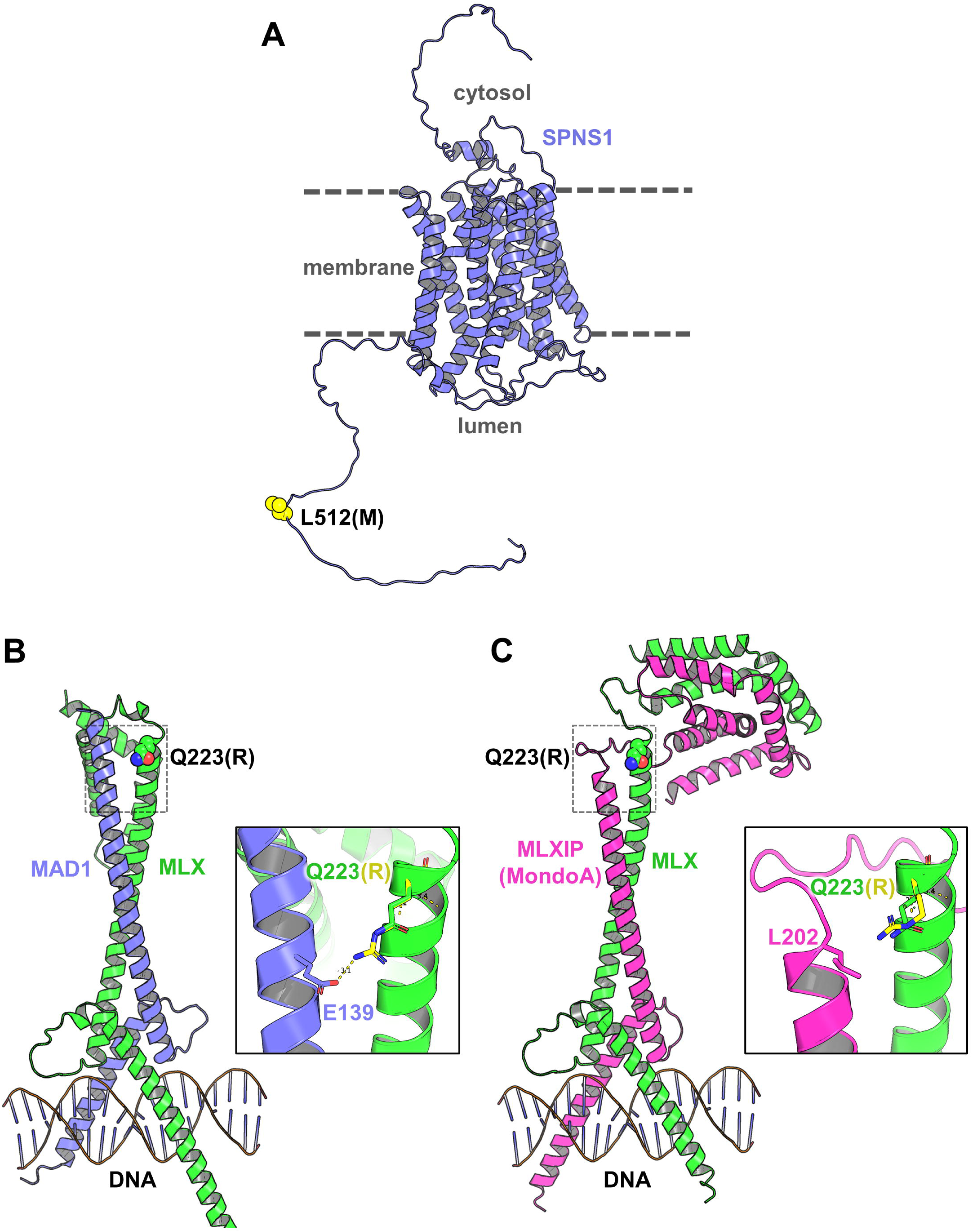
Structural analysis of SPNS1 p.L512M and MLX p.Q223R. A) Alphafold prediction of the structure of the lysophospholipid transporter SPNS1 (alternative isoform, Uniprot #H3BR82). The mutation p.L512M would take place in the lumen of the lysosome. B) AlphaFold model of the MAD1-MLX heterodimer superposed on the structure of the MAD1-MAX-DNA complex (PDB 1NLW). The inset is a zoom on the MLX p.Q223R mutation, displaying the effect that the mutation may have on the interaction with the MAD1 protein. C) AlphaFold model of the MLXIP-MLX heterodimer superposed on the structure of the MAD1-MAX-DNA complex, as described above. Note that AlphaFold also predicts an interaction between the C-termini of MLXIP and MLX (but not MAD1 and MLX).

The Max-like protein (MLX) is at the heart of a transcriptional network pathway involved in energy metabolism and cell signalling.^15,16^ It interacts with at least 6 other related proteins including the MAD family of transcriptional repressors and the Mondo family of transcriptional activators. These proteins contain basic/helix-loop-helix/leucine zipper (bHLHZ) domains that form heterodimers and interact with DNA carrying the CACGTG E-box motif. To understand the impact of the p.Q223R *MLX* mutation (rs665268) on its activity, we modeled the structure of MLX heterodimers with both the MAD and Mondo families using AlphaFold. MLX dimerizes with MAD1,^16^ and thus we superposed its bHLHZ domain on the MAD1-MAX-DNA complex crystal structure^17^ to generate the ternary complex model. The model shows that Gln223 in MLX is at the end of the dimerization “zipper” helix (Figure 5B). The mutation p.Q223R induces the formation of a salt bridge with Glu139 in MAD1, which could strengthen the interaction between MAD1 and MAX. This could then downregulate the interaction of MAD1 with MAX through competition, and thus affect the extent of the transcriptional repression. Glu139 is not conserved in other MAD-related proteins such as MXI1 and MAD3/4. Furthermore, the model of MLX interacting with MLXIP, a protein of the Mondo family also known as MondoA,^18^ shows that the mutation may negatively affect the formation of this heterodimer by introducing a charge next to a hydrophobic sidechain (Figure 5C). The nuclear localization of Mondo proteins is dependent on their interaction with MLX,^15^ and thus the mutation may down regulate activation by the Mondo family while strengthening repression via MAD1.

### Gene enrichment analysis of nominated genes show the inositol phosphate biosynthetic pathway as a novel pathway involved in Parkinson’s disease

We further examined whether the nominated genes highlighted specific pathways and mechanisms associated with PD. We performed a pathway enrichment analysis by examining over-representation of the top nominated genes in biological processes and cellular components using the top genes in each locus. Among the biological processes passing the false discovery rate (FDR) correction, the inositol phosphate biosynthetic process (GO:0032958) and polyol biosynthetic process (GO:0046173) were strongly enriched (Figure 6A). Inositol is associated with 4 candidate genes, namely *ITPKB*, *IP6K2*, *PPIP5K2*, and *INPP5F*. The features most important for nomination of *ITPKB*, *IP6K2*, *PPIP5K2* and *INPP5F* as PD associated genes by our ML model are shown in Figure 4. Cellular components were also identified in the gene enrichment analysis (Figure 6B).

**Figure 6:**
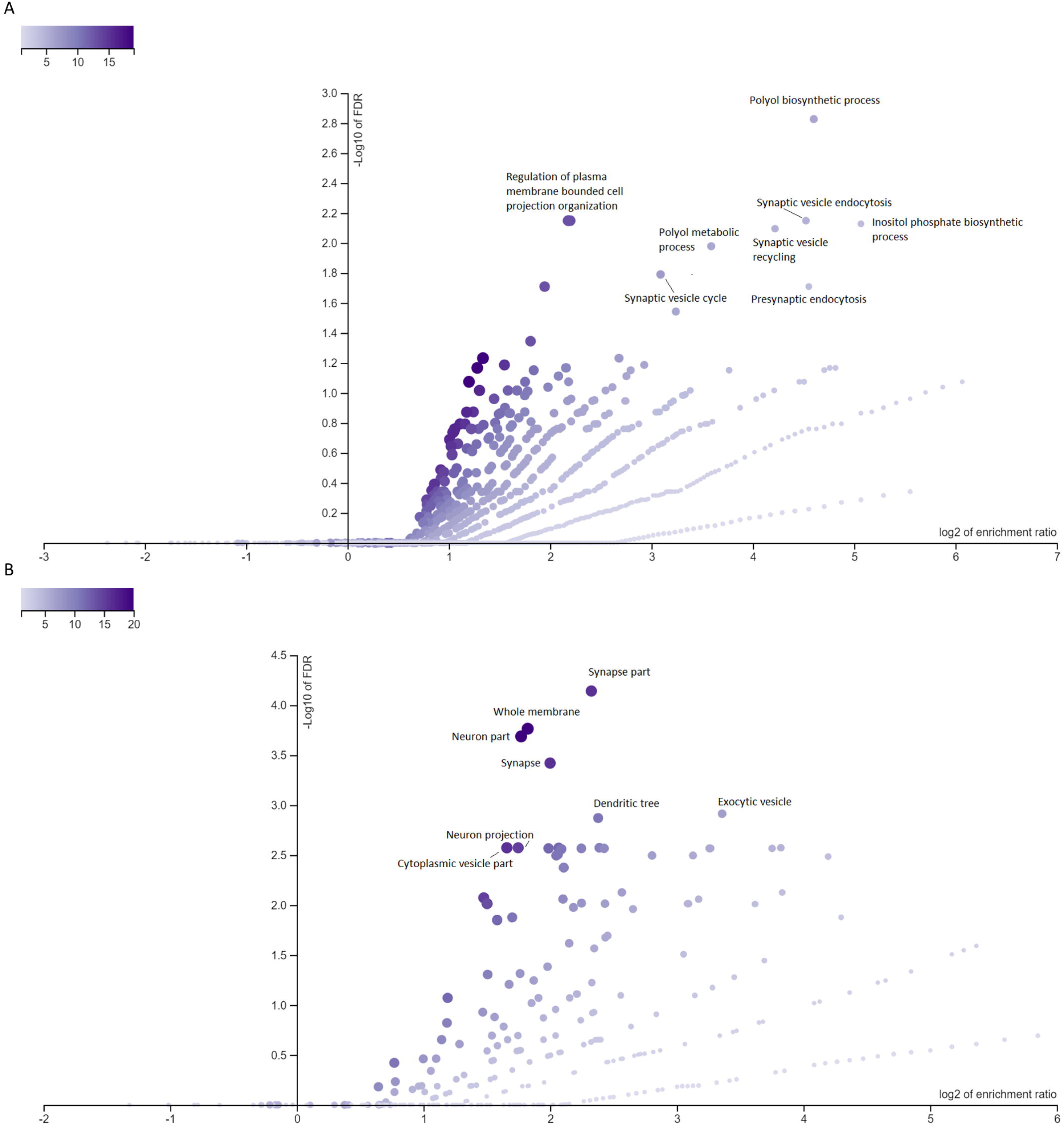
Volcano plots of Gene Ontology biological processes and cellular components. Volcano plots of gene-set enrichment analysis using WebGestalt showing the log of the FDR versus the enrichment ratio. *P*-value are calculated using a hypergeometric test. All pathways that are significant after FDR correction were named.

### Pathway specific polygenic risk score of the inositol phosphate biosynthetic pathway is associated with Parkinson’s disease

To further study the association between the putative novel PD pathways and PD status, pathway-specific polygenic risk scores (PRS) were calculated for the above-mentioned gene sets. The association between these PRS and PD was examined in six PD cohorts, followed by a meta-analysis as detailed in the Methods section. One outlier cohort was excluded due to heterogeneity. The pathway specific PRS were first calculated using all the genes in that pathway. Then, to further validate that the specific pathway is indeed important in PD, we excluded the genes nominated by our machine learning pathway and re-calculated the PRS. By removing these genes with GWAS significant signals, we could examine the residual effect of the remaining of the pathway. The inositol phosphate biosynthetic pathway was associated with PD even after excluding the genes nominated in our analysis (OR 1.06, 95% CI 1.03-1.09, p=7.01E-05), as well as other related pathways (Table 1).

**Table 1:** Meta-analyzes of pathway-specific polygenic risk scores

| Pathway-specific PRS | OR | 95% CI | P | Het P |
| --- | --- | --- | --- | --- |
| POLYOL_BIOSYNTHETIC_PROCESS | 1.20 | 1.17-1.24 | 2.07E-42 | 1.91E-05 |
| INOSITOL_PHOSPHATE_BIOSYNTHETIC_PROCESS | 1.15 | 1.12-1.18 | 2.36E-25 | 1.97E-02 |
| POLYOL_BIOSYNTHETIC_PROCESS_filtered | 1.09 | 1.06-1.12 | 1.04E-09 | 1.12E-02 |
| INOSITOL_PHOSPHATE_BIOSYNTHETIC_PROCESS_filtered | 1.06 | 1.03-1.09 | 1.31E-05 | 1.45E-01 |

PRS: Polygenic risk score, OR: odds ratio, CI: confidence interval; P: p-value, Het: Heterogeneity, filtered: excluded Parkinson's disease top gene, GOBP\_INOSITOL\_PHOSPHATE\_BIOSYNTHETIC\_PROCESS: GeneOntology inositol phosphate biosynthetic process (GO:0032958), GOBP\_POLYOL\_BIOSYNTHETIC\_PROCESS: GeneOntology polyol biosynthetic process (GO:0046173).

### Rare variants in *KCNIP3* and *LSM7* and in the polyol/inositol biosynthetic pathway are associated with Parkinson’s disease

To further establish the potential role of the nominated genes in PD, we performed rare variant burden tests in all the genes nominated by our model. As expected, genes that are known to harbor rare PD coding mutations including *GBA1*, *LRRK2* and *GCH1* were associated with PD (Table 2, Supplementary Table 6). Three additional genes, including two genes that have not been previously implicated in PD (*KCNIP3* and *LSM7*) showed burden of rare variants after FDR correction for multiple comparisons. We then examined the genes from pathway enrichment analysis and found that rare variants in the polyol/inositol biosynthetic pathway were also associated with PD (SKAT-O, p=1.58E-04), further supporting its role in PD.

**Table 2:** Meta-analysis of rare variant analysis of putative causal genes

| Set | P | FDR P |
| --- | --- | --- |
| GBA_Rarefunctional | 2.04E-12 | 6.22E-10 |
| GBA_Rarenonsyn | 3.38E-11 | 5.15E-09 |
| GBA_RareLOF | 1.22E-06 | 1.24E-04 |
| GBA_RareCADD | 2.32E-06 | 1.77E-04 |
| LSM7_RareLOF | 3.69E-06 | 2.25E-04 |
| KCNIP3_RareLOF | 1.12E-05 | 5.69E-04 |
| GCH1_RareLOF | 2.02E-05 | 8.80E-04 |
| LRRK2_RareCADD | 6.07E-05 | 2.31E-03 |
| Polyol_Rarefunctional | 1.59E-04 | 5.38E-03 |
| Polyol_Rarenonsyn | 2.86E-04 | 8.74E-03 |
| NUCKS1_RareCADD | 4.13E-04 | 1.14E-02 |
| Polyol_RareLOF | 1.54E-03 | 3.91E-02 |
| SYT17_Rarenonsyn | 4.61E-03 | 9.37E-02 |
| P2RY12_RareLOF | 4.38E-03 | 9.37E-02 |
| CYLD_RareLOF | 4.48E-03 | 9.37E-02 |
| SYT17_Rarefunctional | 7.39E-03 | 1.38E-01 |
| LCORL_RareLOF | 7.66E-03 | 1.38E-01 |
| CAMK2D_RareLOF | 8.62E-03 | 1.46E-01 |
| FBRSL1_RareLOF | 1.12E-02 | 1.80E-01 |
| CTSB_RareLOF | 1.20E-02 | 1.82E-01 |
| KPNA1_RareCADD | 1.35E-02 | 1.96E-01 |
| ASXL3_RareLOF | 1.52E-02 | 2.10E-01 |
| KPNA1_RareLOF | 1.76E-02 | 2.33E-01 |
| LRRK2_Rarefunctional | 2.57E-02 | 3.14E-01 |
| MICU3_RareLOF | 2.56E-02 | 3.14E-01 |
| VAMP4_Rarenonsyn | 2.93E-02 | 3.43E-01 |
| MBNL2_RareCADD | 3.04E-02 | 3.43E-01 |
| LRRK2_Rarenonsyn | 3.28E-02 | 3.57E-01 |
| KPNA1_Rarefunctional | 3.46E-02 | 3.64E-01 |
| LSM7_Rarefunctional | 3.58E-02 | 3.64E-01 |
| HIP1R_Rarenonsyn | 3.93E-02 | 3.87E-01 |
| KPNA1_Rarenonsyn | 4.23E-02 | 3.91E-01 |
| HIP1R_Rarefunctional | 4.22E-02 | 3.91E-01 |

Set: variant set across genes/pathway, P: p-value, FDR P: false discovery rate p-value, Rarefunctional: rare functional variants, Rarenonsyn: rare nonsynonymous variants, RareLOF: rare loss-of-function variants, RareCADD: rare variants with CADD score above 15.

## Discussion

In this study, we nominated genes that potentially drive the associations with PD for each of the 78 PD GWAS loci, using multi-omic data and machine learning. Our nominated genes include many genes that have not been studied in the context of PD. Additionally, we identified two novel genes with rare variants (*KCNIP3* and *LSM7*) as well as genes with GWAS significant coding variants such as *SPNS1* and *MLX* that could be further studied. Furthermore, our gene enrichment, pathway specific PRS and rare variant analyses strongly support an involvement of the inositol phosphate biosynthetic pathway in PD.

Four genes nominated by our machine learning model belong to the inositol phosphate biosynthetic pathway: *ITPKB*, *IP6K2* and *PPIP5K2* and *SNCA*,^19^ showing a strong enrichment of this pathway. In addition, *INPP5F* is another gene nominated by our analysis that is involved in inositol processing through a parallel pathway.^20^ Our results demonstrate that the inositol pathway-PRS, even when excluding the previously mentioned genes, is still associated with PD. Taken together, our findings support the importance of the inositol phosphate pathway in PD.

Based on the evidence from the candidate inositol genes and previous work on inositol, inositol could potentially be a therapeutic target for PD. In 1999, a clinical trial on inositol was conducted on nine PD patients.^21^ The treatment with inositol compared with placebo did not improve clinical outcomes. However, we cannot rule out inositol and inositol phosphates as potential therapeutic targets as only nine patients were recruited for this trial.

*ITPKB* encodes for a ubiquitous kinase that phosphorylates inositol 1,4,5-trisphosphate (IP3) to inositol 1,3,4,5 tetrakisphosphate (IP4) using a Ca2+/Calmodulin-dependent mechanism. IP3 is a secondary messenger that stimulates calcium release from the endoplasmic reticulum (ER). In primary neurons, *ITPKB* knockdown/overexpression was shown to increase/reduce levels of a-synuclein aggregation.^22^ Additionally, *ITPKB* knockdown in neurons also leads to the accumulation of calcium in mitochondria. This accumulation can impair the process of autophagy, which is crucial for maintaining mitochondrial health. In neuroblastoma cell, *ITPKB* mRNA levels were also shown to be correlated with *SNCA* expression in the cortex and *IPTKB* protein levels were increased in wild a-synuclein, A53T and A30P mutants.^23^ Meanwhile, *IP6K2* and *PPIP5K2* interacts with the same substrates. *IP6K2* converts inositol hexakisphosphate (IP6) to 5-diphosphoinositol pentakisphosphate (5-IP7) or 1-diphosphoinositol pentakisphosphate (1-IP7) to bis-diphosphoinositol tetrakisphosphate (1,5-IP8) while *PPIP5K2* convert 5-IP7 to 1,5-IP8 and IP6 to 1-IP7. ^24^ In mice, *IP6K2* was implicated in cell death, apoptosis and neuroprotection.^25^ One study proposed that *IP6K2* regulates mitophagy via the Parkin/PINK1 pathway, but further evidence would be required to confirm this hypothesis.^25^ *PPIP5K2* has not been previously implicated in PD. It was associated with hearing loss and colorectal carcinoma.^26,27^ Finally, *INPP5F* is involved with a different inositol pathway, it encodes Sac2, which converts phosphoinositides such as PI(4,5)P2 to phosphatidylinositol during endocytosis.^28^

Inositol phosphate has been suggested to be involved in obesity, insulin resistance and energy metabolism.^29^ In postmortem brain tissues, [3H]Inositol 1,4,5-trisphosphate binding sites were found to be reduced in certain brain regions of PD patients such as the caudate nucleus, putamen, and pallidum.^30^ Additionally, IP6 was shown to be associated with PD. IP6 has a neuroprotective effect on dopaminergic cells by preventing 6-OHDA-induced apoptosis.^31^ IP6 inhibits the activity of β-secretase 1 (BACE1), an enzyme that cleaves amyloid-β precursor protein into toxic Aβ peptides.^32^ Paraquet-induced neurodegeneration in *Drosophila* was suggested to increase the levels of inositol phosphates metabolites.^33^ Previous studies have also suggested that different stereoisomers of inositol such as *scyllo*-inositol can inhibit the aggregation of a-synuclein^34^ or decrease of myoinositol concentration in PD patients.^35,36^

Recent studies on inositol investigated the role of *SYNJ1*, an autosomal recessive form of early-onset parkinsonism.^37^ SYNJ1 is a lipid phosphatase of phosphatidylinositol-3,4,5-trisphosphate (PIP3).^38^ SYNJ1 knockout cell models were associated with an increase of a-synuclein and PIP3 levels. PIP3 dysregulation was suggested to promote a-synuclein aggregation, which increase the risk of PD. Together with our data, there is strong evidence for the involvement of the inositol phosphate biosynthetic pathway in PD, and this pathway should be further studied using both basic science and translational approaches.

Outside of the inositol pathway, *SPNS1* and *MLX* were found to be the top causal gene in their respective loci with putative causal missense SNPs: rs7140 and rs665268. Rs7140 corresponds to p.Leu563Val on the *SPNS1* transcript variant X1. We found that *SPNS1* expression is lower in the SOX6_ATGR1 dopaminergic neuron subpopulation in PD compared to controls. This subcluster was previously highlighted to be the most susceptible to neurodegeneration in PD.^11^ *SPNS1* encodes a sphingolipid transmembrane transporter in the lysosome. The autophagy-lysosomal pathway has been well-established to be crucial in PD pathogenesis, especially the lysosomal sphingolipid metabolism pathway, which includes well established PD-associated genes including *GBA1*, *GALC*, *SMPD1* and others.^39,40^ SPNS1 deficiency results in lipid accumulation in the lysosome and impaired lysosomal function.^13^

The second nominated gene in which we identified rare variants, *MLX*, encodes a Max-like protein X which belongs to a family of transcription factors regulating glucose metabolism. Rs665268 is a missense variant (p.Gln139Arg) that was found to be associated with Takayasu’s arteritis, an autoimmune systemic vasculitis.^41^ *MLX* was also reported to be associated with age at onset of Alzheimer’s disease in females.^42^ This variant was suggested to affect two important PD pathways by increasing oxidative stress and suppressing autophagy in immune cells.^41,42^ *SPNS1* and *MLX* have not been previously implicated in PD. These findings indicate that these genes could play a role in PD and should be further studied.

Although we have identified candidate genes and new rare mutations, there are several limitations to this study. The GWAS on which this analysis is based on is only of European populations. Therefore, our results are potentially restricted to Europeans. In addition, the training set for the machine learning model is limited to a small set of known or highly likely PD genes with the assumption of one causal gene per locus. The study also lacked samples for a testing set due to the small number of well-established PD genes. Since these limitations may introduce some bias, we used different strategies such as controlling for an imbalanced dataset and choosing balanced accuracy as an evaluation function to maximize the performance of the model. Although the distance between variants and genes holds significant predictive power in the model, it is crucial to acknowledge that not all top genes can be accurately predicted solely based on distance. Out of the 78 genes analyzed, 13 were not the closest genes in terms of distance from the gene to the top GWAS SNPs, and 25 were not the closest genes based on distance to the transcription start site. Lastly, the meta-analysis of rare variants can also be somewhat biased due to case/control imbalance. Larger GWAS and functional studies will be required to validate our findings.

Our results nominate multiple genes that have not been thoroughly studied in PD and provide foundation for future functional studies of these genes. As larger PD GWASs will nominate more SNPs and loci, prioritizing causal genes will be crucial to understand the underlying biological mechanisms and disease pathophysiology through additional studies. Future gene prioritization studies will also be able to leverage larger datasets with more positive labels as new PD genes get discovered, and therefore increase the accuracy of the predictions.

## Methods

### General design of the study

Our objective was to nominate the most probable genes to be involved in PD from each GWAS locus based on the most recent PD GWAS (see Figure 1 for the study protocol).^1^ To do so, we first defined all the genes and SNPs that are within these loci (see below) and used to a machine learning approach to nominate the top genes in each locus. Based on the previous literature and consensus between authors, we identified seven genes from well-established loci associated with PD that can be considered the likeliest driving genes of their respective loci (*GBA1*, *LRRK2*, *SNCA*, *GCH1*, *MAPT*, *TMEM175*, *VPS13C*). We then acquired data for multiple features, including different distance measures from top SNPs, different QTLs, expression in relevant tissues and cell types and predictions of variant consequences (78 features out of 284 were used after removal of redundant features, Supplementary Table 7). Using the seven well-established PD genes, which were labeled as positive, and 212 genes in the same loci that received negative labels (i.e. not likely to drive the association with PD, since the PD-driving gene is already well-established), we trained a machine learning model. This model enabled us to generate a prediction score for each gene within each locus, assessing their potential involvement in PD. The gene with the highest score in each locus is the nominated gene to be associated with PD. We then performed multiple post hoc analyses to further validate and explore our results: burden tests for rare variants in the top-scoring genes, pathway enrichment and pathway PRS analyses, differential expression analyses and structural analyses for candidate coding variants.

### Definition of loci and genes within each locus

Following the definition by Nalls et al,^1^ all loci were defined based on the 90 independent risk variants (Supplementary Table 2). Variants within 250 kb were merged into a single locus, which led to 78 loci. All protein coding genes within 1 Mb of the risk variants were included in the model. To exclude non-causal variants, echolocatoR was used as a comprehensive fine-mapping model.^43^ This method leverages Bayesian statistical and functional fine-mapping tools as well as epigenomic data to calculate the causal probability of SNPs in a locus.^43^ In our downstream analysis, we incorporated the SNPs nominated by echolocatoR into the credible gene sets generated by the same tool. Furthermore, we included the 90 independent SNPs obtained from the PD GWAS in our analysis.

### Feature preprocessing

To leverage multi-omic data for the machine learning algorithm, we integrated a comprehensive list of datasets (Supplementary Table 7), which included SNP functional annotations, expression and splicing quantitative trait loci (QTL), scRNA, and chromatin interaction. Since distance was previously shown to be the most predictive feature in about 60-70% of GWAS loci, the distance from each SNP to each gene in the locus and the distance to the transcription start site were included in the model.^44^ To predict the severity of variant consequences, we used VEP^10^ and Polyphen-2.^45^ The SNP2GENE function on the FUMA platform was used to perform functional mapping of SNPs to eQTLs.^46^ In the FUMA settings, we chose the UKB release2b 10k European reference panel, a maximum distance of 1000kb from SNPs to gene, and included the MHC region. All other FUMA settings were kept as default. eQTL and 3D chromatin interaction mapping were performed using brain tissues, whole blood, FANTOM and GTEx datasets. Using scRNA datasets from Kamath et al,^11^ we included gene expression from all ten subpopulations of dopaminergic neurons from postmortem brains of 7 PD and 8 control donors. A complete list of all datasets can be found in Supplementary Table 8.

### Neighborhood scores

To integrate the concept of locus and LD in the model, we calculated the neighborhood scores for each feature by transforming the data relative to the best-scoring gene within each locus,^7^ allowing the model to find the highest expressed genes across each locus. For example, if the feature is “maximum gene expression in blood”, the gene with the highest expression in each locus would have a score of one while the score of the remaining genes in the locus would be calculated following the expression of gene divided by the expression of highest expressed gene in the locus. Negative log transformation was applied so that the closest gene had the highest score.

### Machine learning model to prioritize genes

We used XGBoost^47^ to train the machine learning model. We selected well-established genes from PD loci for the training dataset (*GBA1*, *GCH1*, *LRRK2*, *MAPT*, *SNCA*, *TMEM175*, *VPS13C*). These genes were labeled as positive labels, and the remaining genes from these same loci were labeled as negative labels. In total, the training set was composed of 212 genes (7 positive labeled and 205 negative labeled). The scale_pos_weight parameter in XGBoost was set to the ratio of negative to positive labels to control for the imbalance. The training process involved two steps. Firstly, the model was trained to identify redundant features, eliminating any redundant or uninformative variables from the dataset. In the second step, the final training model was created using the selected features. This two-step approach helps optimize the training process and ensures that the model focuses on relevant and informative features to make accurate predictions. We performed hyperparameter tuning and five-fold cross-validation on both models. Mean average precision was used as an evaluation function to maximize the correct positive predictions made. Of the 284 features, 78 features passed feature selection for the final training model.

### Functional enrichment analysis

To examine whether specific pathways may be involved in PD, based on the genes nominated in each locus, we performed an over-representation analysis using WebGestalt (WEB-based GEne SeT AnaLysis Toolkit) on January 25, 2023.^48^ We included the top candidate gene from each locus, and examined enrichment in terms of biological processes and cellular components from the Gene Ontology data. The genome protein-coding list was used as the reference list and pathways were considered to be associated with PD if significant after FDR correction.

### Single-cell and bulk RNAseq analyses

To examine whether genes nominated by the machine learning model may be differentially expressed in PD relevant models, we used publicly available single-cell and bulk RNAseq data from FOUNDIN-PD^12^ and Kamath et al.^11^ FOUNDIN-PD scRNA data includes 80 induced pluripotent stem cell (iPSC) lines collected after 65 days.^12^ We then performed differential gene expression analyses between PD cases and controls. For scRNA, we used the MAST^49^ package after adjusting for covariates such as age, sex and batch. For bulk RNAseq, we used DESeq2^50^ while adjusting for the same covariates.

### Pathway polygenic risk score analyses

Pathway-specific PRS analysis can further support a role for specific pathways in PD.^51^ Using PRSet,^52^ pathway-specific polygenic risk scores (PRS) were calculated for pathways nominated by gene set analysis on 14,828 PD cases and 13,283 controls from seven cohorts (McGill, Parkinson’s Progression Markers Initiative (PPMI), Vance (dbGap phs000394), International Parkinson’s Disease Genomics Consortium (IPDGC) NeuroX dataset (dbGap phs000918.v1.p1), National Institute of Neurological Disorders and Stroke (NINDS) Genome-Wide genotyping in Parkinson’s Disease (dbGap phs000089.v4.p2), NeuroGenetics Research Consortium (NGRC) (dbGap phs000196.v3.p1) and UK Biobank). The number of cases and control for each cohort is described in Supplementary Table 9. Participants were unrelated individuals of European ancestry and were not gender matched. Rare SNPs (minor allele frequency < 0.01) with p-value < 0.05 were excluded from the analysis. LD clumping was performed using r2=0.1 and 250kb distance. A permutation testing was performed with 10 000 label permutations to generate empirical p-value for each gene set after adjusting for a prevalence of 0.005, age at onset for cases, age at enrollment for control, sex, and the top 10 principal components. The Vance cohort was excluded from the meta-analysis due to significant heterogeneity.

### Rare variant burden analyses

To examine whether there is an association between rare variants in the genes nominated by the machine learning model and PD, we used MetaSKAT^53^ to perform a meta-analyses of rare variants. We used whole exome sequencing (WES) available for 602 PD patients, 6,284 proxy patients and 140,207 controls from UK Biobank (n=147,093) and 2,600 PD patients, 3,677 controls from Accelerating Medicines Partnership Parkinson’s Disease (AMP-PD)^54^ datasets (n=6277). Additional selection criteria for UK Biobank and AMP PD were reported previously.^55,56^ We performed the analysis on several groups of rare variants (allele frequency < 0.01): loss of function variants, nonsynonymous variants, potentially deleterious (CADD>20) variants and functional (including nonsynonymous, frame-shift, stop-gain, and splicing) variants. Pathway-specific rare variant analysis was performed by combining PD genes from the pathways nominated previously. All analyses were adjusted for age at onset for cases, age at sample for control and sex.

### Structural analysis

The atomic coordinates of SPNS1 (Uniprot #H3BR82) were retrieved from the AlphaFold server (https://alphafold.ebi.ac.uk/). The structures of MLX-MAD1 and MLX-MLXIP were generated using AlphaFold-Multimer version 3, as implemented in ColabFold.^57,58^ The ternary complex with a DNA duplex was generated by superposing the heterodimers on the crystal structure of the MAD1-MAX-DNA complex (PDB 1NLW). The figures were generated using PyMol v.2.4.0.

## Data availability

The data used for this study can be accessed on: FUMA https://fuma.ctglab.nl/; Cuomo et al. 2020; Bryois et al. 2021; PPMI ppmi-info.org; UK Biobank https://www.ukbiobank.ac.uk/, SMR https://yanglab.westlake.edu.cn/software/smr/; and Kamath et al 2021.

## Code availability

The scripts used for this study can be found on GitHub: github.com/gan-orlab/gene_prio

## Data Availability

https://ppmi-info.org

https://www.ukbiobank.ac.uk/

https://yanglab.westlake.edu.cn/software/smr/

## Acknowledgement

This work was financially supported by the Michael J. Fox Foundation, Parkinson’s Society Canada, the Canadian Consortium on Neurodegeneration in Aging (CCNA), Canadian Institutes of Health Research (#476751), and the Canada First Research Excellence Fund (CFREF) awarded to McGill University for the Healthy Brains Healthy Lives (HBHL) program. S.R. received a scholarship from the Canadian Institutes of Health Research. Z.G.O. is supported by the Fonds de recherche du Quebec Sante (FRQS) Chercheurs boursiers award and is a William Dawson and Killam Scholar.

UK Biobank Resources were accessed under application number 45551. Data used in the preparation of this article were obtained from the AMP PD Knowledge Platform. For up-to-date information on the study, visit https://www.amp-pd.org. The AMP® PD program is a public-private partnership managed by the Foundation for the National Institutes of Health and funded by the National Institute of Neurological Disorders and Stroke (NINDS) in partnership with the Aligning Science Across Parkinson’s (ASAP) initiative; Celgene Corporation, a subsidiary of Bristol-Myers Squibb Company; GlaxoSmithKline plc (GSK); The Michael J. Fox Foundation for Parkinson’s Research; Pfizer Inc.; Sanofi US Services Inc.; and Verily Life Sciences.

Clinical data and biosamples used in preparation of this article were obtained from the (i) Michael J. Fox Foundation for Parkinson’s Research (MJFF) and National Institutes of Neurological Disorders and Stroke (NINDS) BioFIND study, (ii) Harvard Biomarkers Study (HBS), (iii) NINDS Parkinson’s Disease Biomarkers Program (PDBP), (iv) MJFF Parkinson’s Progression Markers Initiative (PPMI), and (vii) NINDS Study of Isradipine as a Disease-modifying Agent in Subjects With Early Parkinson Disease, Phase 3 (STEADY-PD3).

BioFIND is sponsored by The Michael J. Fox Foundation for Parkinson’s Research (MJFF) with support from the National Institute for Neurological Disorders and Stroke (NINDS). The BioFIND Investigators have not participated in reviewing the data analysis or content of the manuscript. For up-to-date information on the study, visit michaeljfox.org/news/biofind.

The Harvard Biomarker Study (HBS) is a collaboration of HBS investigators [full list of HBS investigators found at https://www.bwhparkinsoncenter.org/biobank/ and funded through philanthropy and NIH and Non-NIH funding sources. The HBS Investigators have not participated in reviewing the data analysis or content of the manuscript.

PPMI is sponsored by The Michael J. Fox Foundation for Parkinson’s Research and supported by a consortium of scientific partners: [list the full names of all of the PPMI funding partners found at https://www.ppmi-info.org/about-ppmi/who-we-are/study-sponsors]. The PPMI investigators have not participated in reviewing the data analysis or content of the manuscript. For up-to-date information on the study, visit www.ppmi-info.org.

The Parkinson’s Disease Biomarker Program (PDBP) consortium is supported by the National Institute of Neurological Disorders and Stroke (NINDS) at the National Institutes of Health. A full list of PDBP investigators can be found at https://pdbp.ninds.nih.gov/policy. The PDBP investigators have not participated in reviewing the data analysis or content of the manuscript.

The Study of Isradipine as a Disease-modifying Agent in Subjects With Early Parkinson Disease, Phase 3 (STEADY-PD3) is funded by the National Institute of Neurological Disorders and Stroke (NINDS) at the National Institutes of Health with support from The Michael J. Fox Foundation and the Parkinson Study Group. For additional study information, visit https://clinicaltrials.gov/ct2/show/study/NCT02168842. The STEADY-PD3 investigators have not participated in reviewing the data analysis or content of the manuscript.

## Authors contributions

E.Y. contributed to conception and design of the study; acquisition, analysis and interpretation of data; drafted and substantively revised the manuscript. R.L. contributed to design of the study and substantively revised the manuscript. R.A.T contributed to design of the study and substantively revised the manuscript. L.L. contributed to the acquisition, analysis and interpretation of data and substantively revised the manuscript. K.S. contributed to the acquisition, analysis and interpretation of data and substantively revised the manuscript. S.R. substantively revised the manuscript. J-F.T. contributed to the acquisition, analysis and interpretation of data and substantively revised the manuscript. E.A.F. contributed to design of the study and substantively revised the manuscript. Z.G-O. contributed to conception and design of the study and substantively revised the manuscript.

