## Supplemental Figure for "Identification of novel variants, genes and pathways potentially linked to Parkinson’s disease using machine learning"

**A**
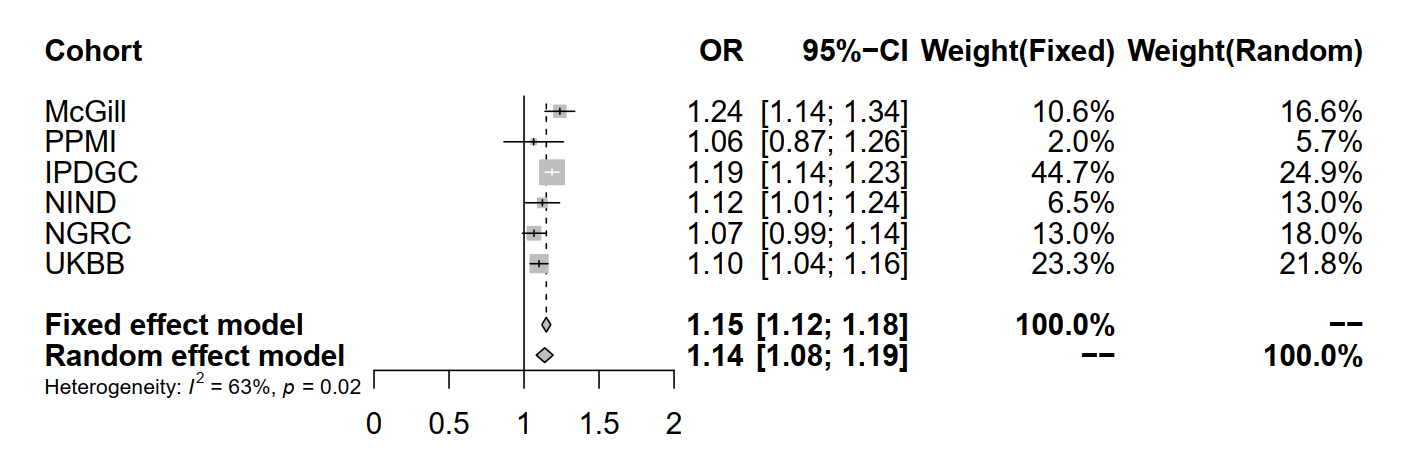
**B**
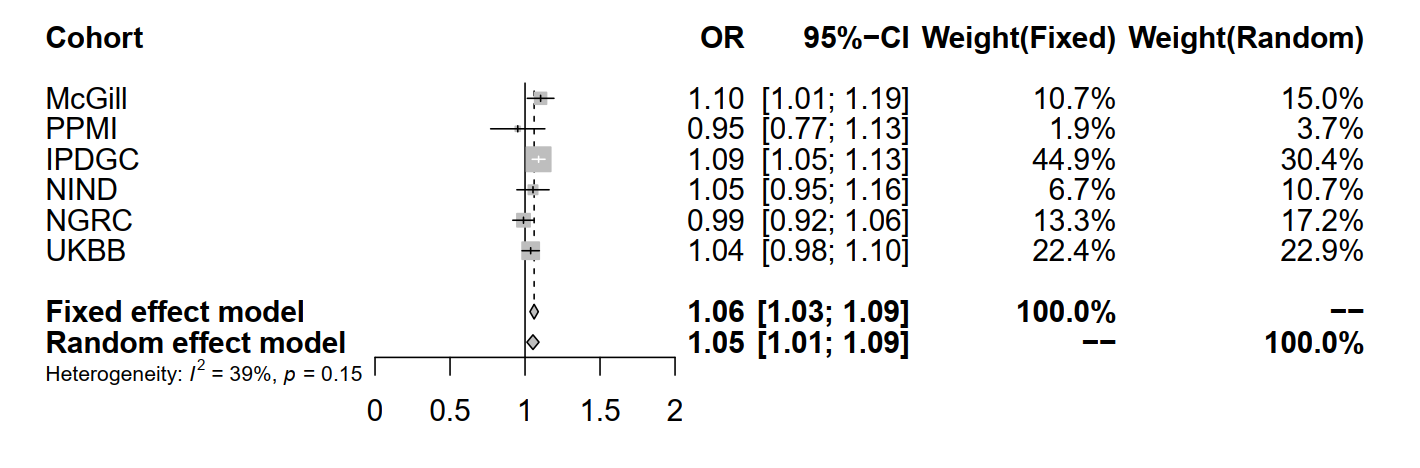
**C**
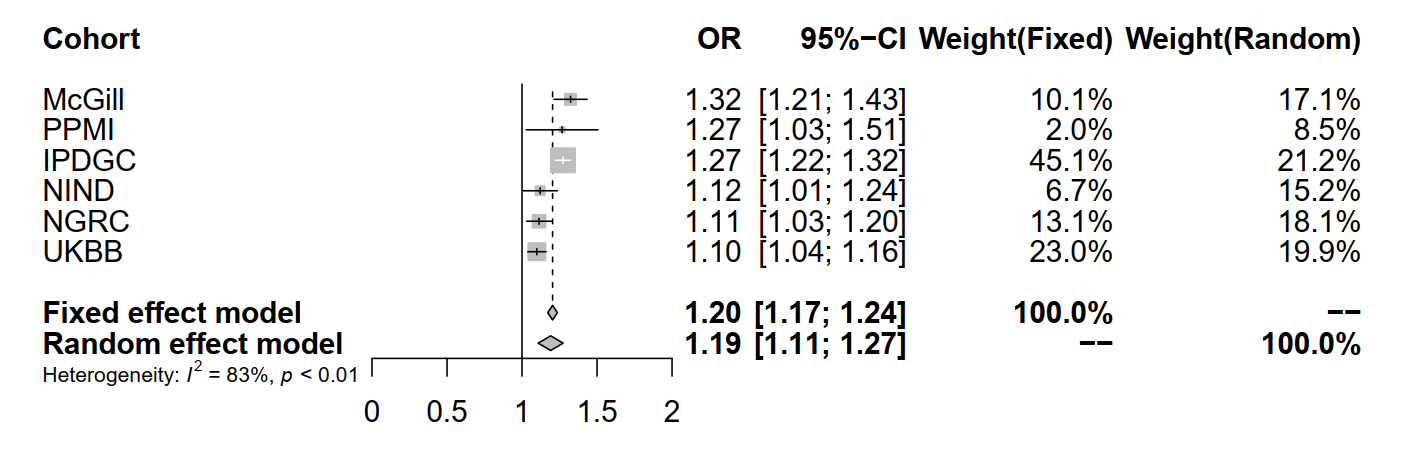
**D**
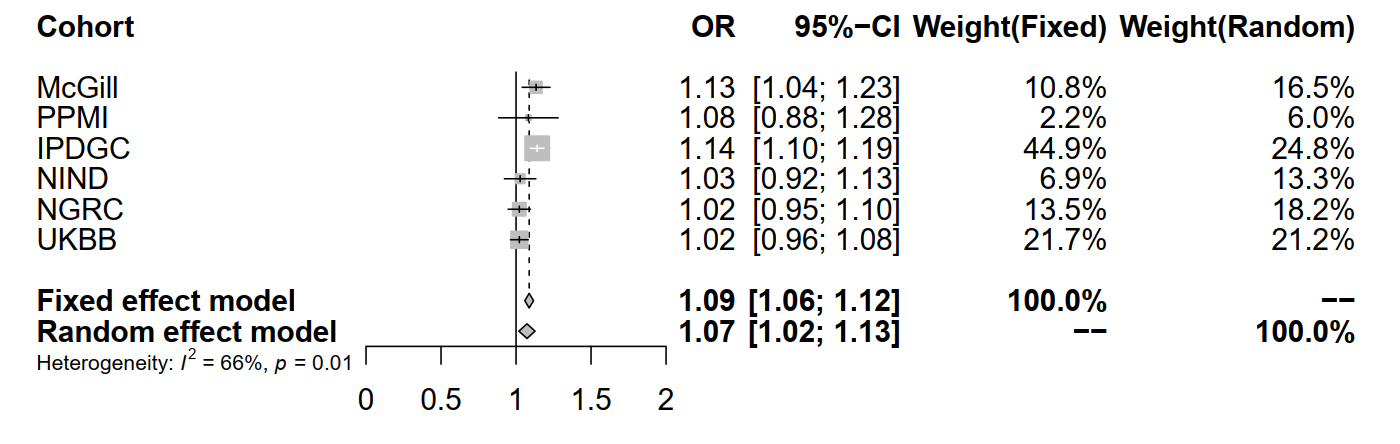

**Supplementary Figure S1: Forest plot of meta-analysis of pathway-specific polygenic risk score.** A) Inositol phosphate biosynthetic process (GO:0032958) B) Inositol phosphate biosynthetic process (GO:0032958) excluding Parkinson’s disease GWAS loci candidate gene C) Polyol biosynthetic process (GO:0046173) D) Polyol biosynthetic process (GO:0046173) excluding Parkinson’s disease GWAS loci candidate gene
